# Uptake of same-day initiation of HIV treatment in Malawi, South Africa, and Zambia: the SPRINT retrospective cohort study

**DOI:** 10.1101/2022.11.28.22282854

**Authors:** Amy Huber, Kamban Hirasen, Alana T Brennan, Bevis Phiri, Timothy Tchereni, Lloyd Mulenga, Prudence Haimbe, Hilda Shakwelele, Rose Nyirenda, Bilaal Wilson Matola, Andrews Gunda, Sydney Rosen

## Abstract

**Introduction:** Since 2017 global guidelines have recommended “same-day initiation” (SDI) of antiretroviral treatment (ART) for patients considered ready for treatment on the day of HIV diagnosis. Many countries in sub-Saharan Africa have incorporated a SDI option into national guidelines, but uptake of SDI is not well documented. We estimated average time to ART initiation at 12 public healthcare facilities in Malawi, 5 in South Africa, and 12 in Zambia.

**Methods:** We sequentially enrolled patients who were eligible to start ART between January 2018 and June 2019 and reviewed their medical records from the point of HIV treatment eligibility (HIV diagnosis or first HIV-related interaction with the clinic) to the earlier of treatment initiation or 6 months. We estimated the proportion of patients initiating ART at their original healthcare facilities on the same day or within 7, 14, 30, or 180 days of baseline, stratified by country and gender.

**Results:** We enrolled 826 patients in Malawi, 534 in South Africa, and 1,984 in Zambia. 88% of patients in Malawi, 57% in South Africa, and 91% in Zambia were offered and accepted SDI. In Malawi, most patients who did not receive SDI had also not initiated ART ≤ 6 months. In South Africa, an additional 13% of patients initiated ≤ 1 week, but 21% still had no record of initiation ≤ 6 months. Among those who did initiate within 6 months in Zambia, most started ≤ 1 week. There were no major differences by gender. Both WHO Stage III/IV and tuberculosis symptoms were associated with delays in ART initiation.

**Discussion:** As of 2020, uptake of same-day ART initiation was widespread, if not nearly universal, in Malawi and Zambia but was considerably less common in South Africa. Limitations of the study include pre-COVID-19 data that do not reflect pandemic adaptations and potentially missing data for Zambia. South Africa may be able to increase overall ART coverage by reducing numbers of patients who do not initiate ≤ 6 months.

**Registration:** Clinicaltrials.gov NCT04468399 (Malawi), NCT04170374 (South Africa), and NCT04470011 (Zambia).

## INTRODUCTION

In 2017, the World Health Organization (WHO) began recommending “same-day initiation” (SDI) of treatment for HIV patients considered clinically and personally ready for antiretroviral therapy (ART)[1]. This recommendation was based on a series of randomized trials and observational studies that had demonstrated improved retention in care and viral suppression rates for patients offered SDI, compared to those offered what was then the standard of care, which typically required multiple clinic visits before a patient received an initial supply of antiretroviral medications (ARVs)[2]. In the clinical trials, most of the improvement arose from a reduction in loss to follow up between testing positive for HIV and initiating ART. Observational studies have reported that among patients actually starting ART on the same day under routine care, loss to follow up during the first year after initiation has tended to be higher than in the pre-same day initiation period[3,4], but these studies have generally compared outcomes among patients who started on the same day to outcomes among those known to have started within a specified number of days, ignoring patients who never started ART at all.

In response to the WHO recommendation, many countries adopted SDI into their national guidelines. Instructions to healthcare facilities as to exactly how to implement SDI and to which patients it should be offered, however, were not precise, allowing the interval between a patient’s first clinic visit and ART initiation to be determined by provider judgment and/or patient preference. Time intervals from a patient’s initial presentation (HIV diagnosis or first HIV-related healthcare interaction) to initiation of ART are not routinely reported in electronic medical record (EMR) systems, which typically only create a record for a patient starting at the time of ART initiation. Little evidence is available on the proportion of patients actually starting ART on the same day or the characteristics of those who do or do not. Without this information, it is difficult to understand observed differences or identify opportunities for improving care.

This paper reports quantitative results of the SPRINT study (Survey of Procedures and Resources for Initiating Treatment of HIV in Africa). It documents uptake of SDI, time to ART initiation for those not accepting SDI, and predictors of SDI in three high HIV burden countries in sub-Saharan Africa.

## METHODS

### Study sites

SPRINT was a retrospective record review conducted in Malawi, South Africa, and Zambia. In each country, in collaboration with national health officials, we first purposively selected provinces and districts that provided diversity of setting (rural/urban) and of nongovernmental supporting partner organizations. We then identified a convenience sample of public sector primary healthcare clinics within these provinces or districts and conducted visits to these sites to assess facility-level and HIV testing and initiation-specific aggregate indicators. The final sites selected provided diversity in size and geographic setting, had relatively strong routine data collection, and were welcoming of the research team. Selected study sites are described in Table 1.

**Table 1.**
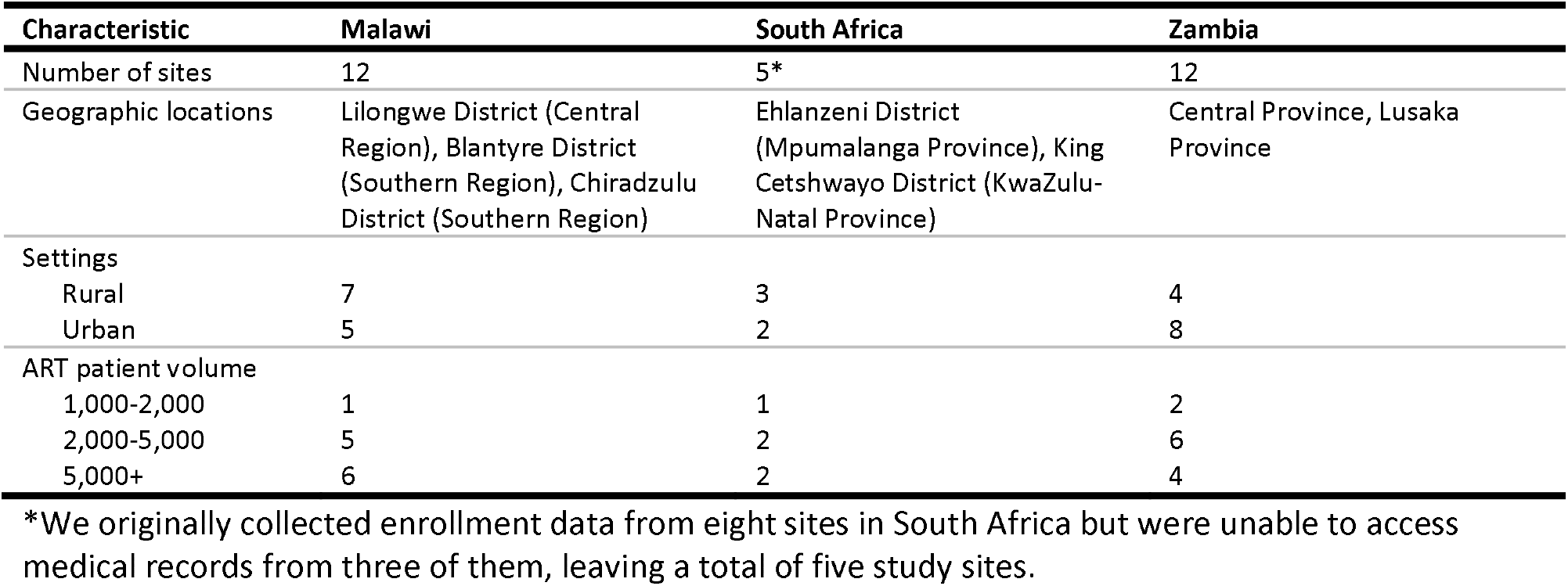
SPRINT study sites.

| Characteristic | Malawi | South Africa | Zambia |
| --- | --- | --- | --- |
| Number of sites | 12 | 5* | 12 |
| Geographic locations | Lilongwe District (Central Region), Blantyre District (Southern Region), Chiradzulu District (Southern Region) | Ehlanzeni District (Mpumalanga Province), King Cetshwayo District (KwaZulu-Natal Province) | Central Province, Lusaka Province |
| Settings |  |  |  |
| Rural | 7 | 3 | 4 |
| Urban | 5 | 2 | 8 |
| ART patient volume |  |  |  |
| 1,000-2,000 | 1 | 1 | 2 |
| 2,000-5,000 | 5 | 2 | 6 |
| 5,000+ | 6 | 2 | 4 |

\*We originally collected enrollment data from eight sites in South Africa but were unable to access medical records from three of them, leaving a total of five study sites.

At the time of study enrollment, guidelines for ART initiation in the three study countries encouraged but did not require that same-day initiation be offered to patients whom providers determined to be eligible, without waiting for CD4 count results. Tuberculosis (TB) symptom screening was recommended in all three countries, with a TB test conducted for symptomatic patients prior to ART initiation. Guidelines did not specify how long to delay ART initiation among patients diagnosed with active TB but generally encouraged starting HIV treatment as soon as possible[5–7].

### Enrollment and data collection

Patients were eligible for enrollment in the study if they were 1) non-pregnant adults (≥ 18 years old); 2) tested HIV-positive or provided documentation of HIV infection (prior positive test); and 3) were eligible to start or re-start ART at one of the study sites between 01 January to 31 December 2018 in South Africa or between 01 July 2018 and 30 June 2019 in Malawi and Zambia. We sequentially enrolled up to 200 patients per site, starting on the latest date in the enrollment period and working backward in time until the target sample size was reached. Study-eligible patients were selected from the register, database, or files kept by the site. Medical record data were downloaded from EMRs where available. If an enrolled patient was not found in the EMR or if an EMR entry was incomplete, we manually accessed individual clinic files and/or other clinic registers or databases.

Records were reviewed from the point of HIV treatment eligibility (HIV diagnosis or first interaction with the clinic) to the point of treatment initiation or for a 6-month period, whichever was shorter. By starting with eligibility for treatment, rather than with treatment initiation, the study captured information about patients who failed to initiate treatment within 6 months (dropped out of care before being dispensed medications) and allowed us to observe services received and days elapsed before initiation.

### Sample size, outcomes, and statistical analysis

Study enrollment numbers were determined by resource availability. Our maximum sample size was 200 patients per site, or up to a total of 2,400 patients each in Malawi and Zambia and 1,600 in South Africa. The primary outcome of interest for the study was SDI, defined as being dispensed an initial supply of ARV medications on the day of testing positive for HIV or the day of first HIV-related clinic interaction, if a positive test had been conducted previously at another location, without subsequent linkage to care.

Simple descriptive statistics were used to describe the study sample. Predictors of SDI were assessed using modified Poisson regression[8], clustered by study site to allow intragroup correlation in the standard errors. For our first model (Model 1) including data from all three countries, we controlled for biological sex, age (categorized as 18-29, 30-39, 40-49 and >50 years of age), WHO stage III/IV (vs. I/II) and country, all assessed at ART initiation, as potential risk factors for SDI. Considering that there were no CD4 cell count or TB symptom data from Malawi, we then ran the same model (Model 2) restricted to Zambia and South Africa and included a variable indicating whether the patient received a CD4 count (yes/no) and the presence of TB symptoms (yes/no) at initiation.

### Ethics

Ethical clearance was provided by the Boston University Medical Campus Institutional Review Board (IRB) (Ref No. H-40354 (Malawi), H-39330 (South Africa) and H-40488 (Zambia)), the University of the Witwatersrand Human Ethics Research Committee in South Africa (Ref No. M200238 (Malawi), M190745 (South Africa) and M200599 (Zambia)), the National Health Sciences Research Committee in Malawi (Ref No. 20/04/2458) and the ERES Converge Institutional Review Board in Zambia (Ref No. 2020-Feb-009). The Ministry or Department of Health in all three countries and the National Health Research Authority in Zambia also approved the study. As this was a retrospective review of medical records and no identifiers were collected, informed consent was not sought.

## RESULTS

Between 02 June 2018 and 16 December 2020, we enrolled a total 3,374 participants. After excluding 30 participants who were found to be ineligible (under 18 years old or initiated treatment outside the study period), the final analytic sample included 3,344 participants (826 patients in Malawi, 534 in South Africa, and 1984 in Zambia). CD4 counts and TB status were not available for patients in Malawi. There were no important differences in patient characteristics among the three countries, as shown in Table 1.

### Time to ART initiation

Time to ART initiation is presented in Table 3. Same-day initiation was the norm in Malawi and Zambia, where a vast majority of patients did start ART on the day of diagnosis or first HIV-related clinic interaction. It was much less common in South Africa, where a third of patients had not initiated ART at all within the 6-month study observation period. Among patients who did not initiate on the same day, varying proportions proceeded to initiate within 1, 2, 4, and 24 weeks, as shown in Table 3.

Across all three countries, as shown in Tables 2 and 3, we identified a total of 182 participants who did not initiate treatment within 6 months of HIV diagnosis or their first recorded, HIV-related clinic visit (non-initiators). These patients comprised 8% of the cohort in Malawi, 21% in South Africa, and just 0.2% in Zambia. We note that while these proportions are consistent with expectations in Malawi and South Africa, it is likely that non-initiators in Zambia who should have been included in our sample were not. Although we made multiple attempts to confirm the completeness of our cohort in Zambia, we believe it is likely that more than 0.2% of patients in Zambia who were eligible to initiate ART did not start within 6 months. Among those who did not initiate within 6 months, in Malawi most either died or remained alive and in care, but not on ART, while in South Africa most had died or were simply missing follow up records (Table 3).

**Table 2.**
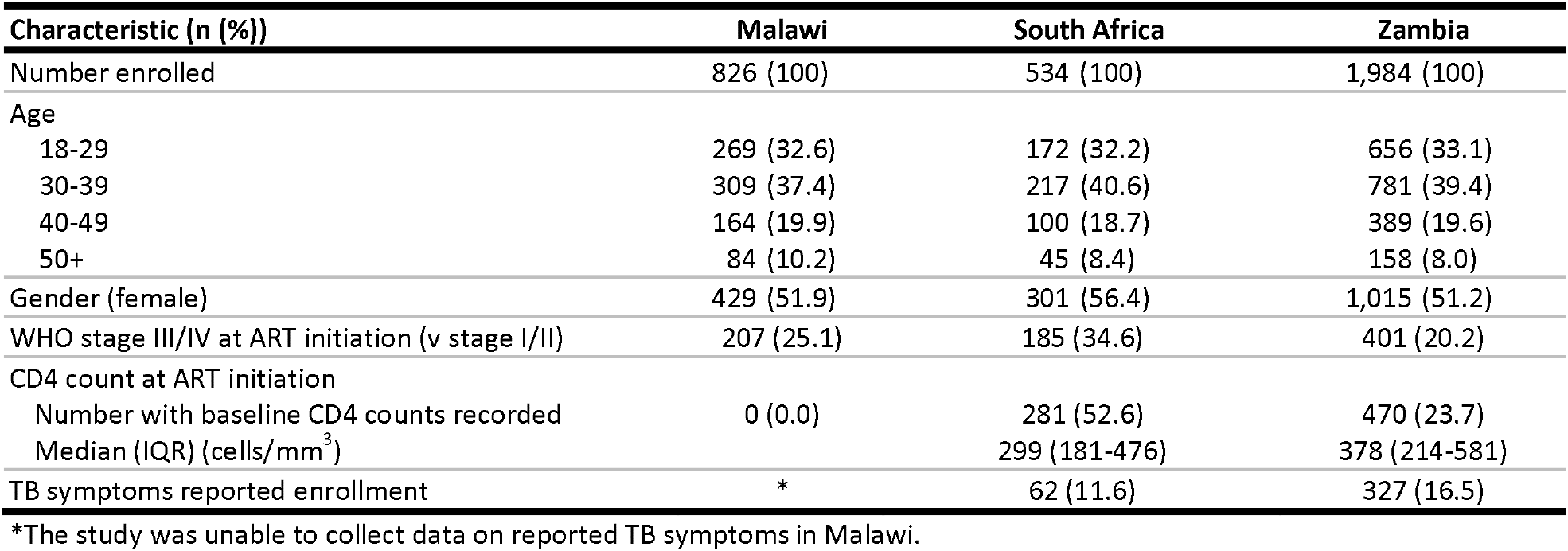
Characteristics of the analytic study sample (n=3,344)

**Table 3.**
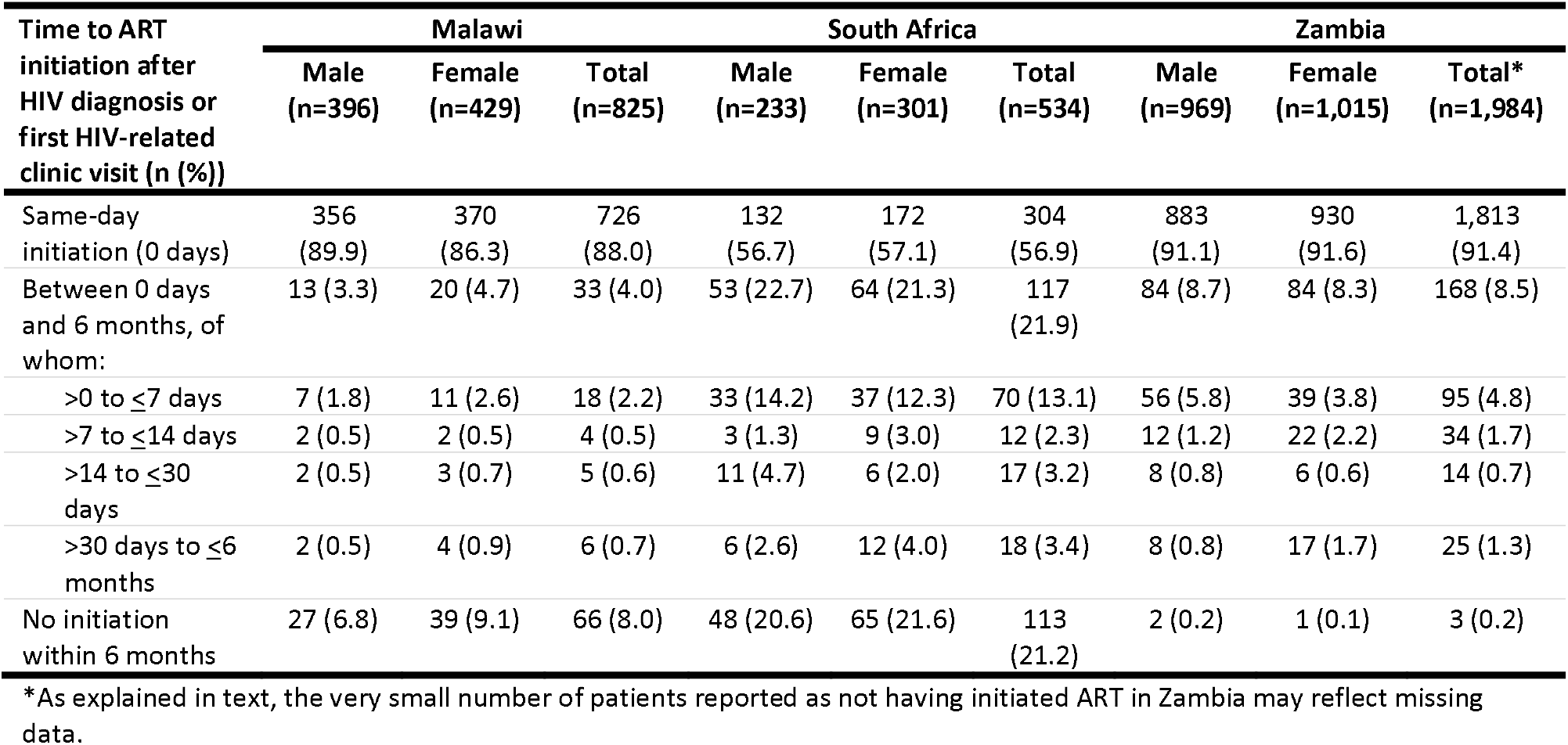
Time to ART initiation (n=3,344)

| Time to ART initiation after HIV diagnosis or first HIV-related clinic visit (n (%)) | Malawi |  |  | South Africa |  |  | Zambia |  |  |
| --- | --- | --- | --- | --- | --- | --- | --- | --- | --- |
|  | Male (n=396) | Female (n=429) | Total (n=825) | Male (n=233) | Female (n=301) | Total (n=534) | Male (n=969) | Female (n=1,015) | Total* (n=1,984) |
| Same-day initiation (0 days) | 356 (89.9) | 370 (86.3) | 726 (88.0) | 132 (56.7) | 172 (57.1) | 304 (56.9) | 883 (91.1) | 930 (91.6) | 1,813 (91.4) |
| Between 0 days and 6 months, of whom: | 13 (3.3) | 20 (4.7) | 33 (4.0) | 53 (22.7) | 64 (21.3) | 117 (21.9) | 84 (8.7) | 84 (8.3) | 168 (8.5) |
| >0 to ≤7 days | 7 (1.8) | 11 (2.6) | 18 (2.2) | 33 (14.2) | 37 (12.3) | 70 (13.1) | 56 (5.8) | 39 (3.8) | 95 (4.8) |
| >7 to ≤14 days | 2 (0.5) | 2 (0.5) | 4 (0.5) | 3 (1.3) | 9 (3.0) | 12 (2.3) | 12 (1.2) | 22 (2.2) | 34 (1.7) |
| >14 to ≤30 days | 2 (0.5) | 3 (0.7) | 5 (0.6) | 11 (4.7) | 6 (2.0) | 17 (3.2) | 8 (0.8) | 6 (0.6) | 14 (0.7) |
| >30 days to ≤6 months | 2 (0.5) | 4 (0.9) | 6 (0.7) | 6 (2.6) | 12 (4.0) | 18 (3.4) | 8 (0.8) | 17 (1.7) | 25 (1.3) |
| No initiation within 6 months | 27 (6.8) | 39 (9.1) | 66 (8.0) | 48 (20.6) | 65 (21.6) | 113 (21.2) | 2 (0.2) | 1 (0.1) | 3 (0.2) |

\*As explained in text, the very small number of patients reported as not having initiated ART in Zambia may reflect missing data.

### Predictors of same-day initiation

Table 4 presents the results of the modified Poisson regression. In Model 1, which included data from all three countries, we found that patients with a WHO stage III/IV (vs. stage I/II) had a 34% decrease in the likelihood of SDI (adjusted risk ratio (aRR): 0.66; 95% confidence interval (CI): 0.54-0.81). Patients in South Africa had a 28% decrease in the likelihood of SDI (aRR: 0.72; 95% CI: 0.63-0.82) compared to Zambia, while the uptake of SDI in Malawi was comparable to that of Zambia. We did not control for CD4 counts or TB symptoms in this model, as these were not available in Malawi.

**Table 4.** Outcomes for those who did not initiate (n=182)

| 6-month outcome for those who did not initiate within 6 months (n (%)) | Malawi<br>n=66 | South Africa<br>n=113 | Zambia<br>n=3 | Total<br>n=182 |
| --- | --- | --- | --- | --- |
| Alive and in care | 20 (30.3) | 4 (3.5) | - | 24 (13.2) |
| Lost to follow up | 1 (1.5) | 1 (0.9) | - | 2 (1.1) |
| Died | 38 (57.6) | 44 (38.9) | - | 82 (45.1) |
| Transferred out | 4 (6.1) | 15 (13.3) | - | 19 (10.4) |
| Missing (no follow up record) | 3 (4.6) | 49 (43.4) | 3 (100) | 55 (30.2) |

**Table 5.** Crude and adjusted models clustered by study site for the outcome of SDI for the entire sample (Model 1) and for South Africa and Zambia where CD4 counts and TB data were collected (Model 2)

| Characteristic | Measure | Model 1<br>SDI (n=3,344) |  | Model 2<br>SDI (n=2,518)* |  |
| --- | --- | --- | --- | --- | --- |
|  |  | Crude RR<br>(95% CI) | Adjusted RR<br>(95% CI) | Crude RR<br>(95% CI) | Adjusted RR (95%<br>CI) |
| Sex | Female | ref. | ref. | ref. | ref. |
|  | Male | 1.02 (0.98-1.05) | 1.01 (0.99-1.03) | 1.01 (0.97-1.05) | 1.00 (0.98-1.03) |
| Age (years) | 18-29.9 | ref. | ref. | ref. | ref. |
|  | 30-39.9 | 1.00 (0.95-1.05) | 1.00 (0.96-1.03) | 1.00 (0.94-1.06) | 0.99 (0.95-1.04) |
|  | 40-49.9 | 1.01 (0.96-1.07) | 0.99 (0.95-1.02) | 1.01 (0.94-1.09) | 0.99 (0.95-1.03) |
|  | ≥50 | 1.00 (0.96-1.05) | 0.99 (0.96-1.03) | 1.02 (0.97-1.08) | 1.01 (0.97-1.06) |
| WHO stage | I/II | ref. | ref. | ref. | ref. |
|  | III/IV | 0.64 (0.51-0.80) | 0.66 (0.54-0.81) | 0.62 (0.46-0.83) | 0.66 (0.52-0.85) |
| Country | Zambia | ref. | ref. | ref. | ref. |
|  | Malawi | 0.96 (0.88-1.05) | 0.98 (0.91-1.05) |  |  |
|  | South Africa | 0.62 (0.47-0.82) | 0.72 (0.63-0.82) | 0.62 (0.47-0.83) | 0.69 (0.60-0.78) |
| TB symptoms | no |  |  | ref. | ref. |
|  | yes |  |  | 0.83 (0.70-0.98) | 0.84 (0.73-0.96) |
| CD4 count done | no |  |  | ref. | ref. |
|  | yes |  |  | 1.07 (0.96-1.19) | 1.09 (0.97-1.22) |

\*Restricted to South Africa and Zambia only

In Model 2, restricted to South Africa and Zambia, we found similar results to Model 1. Patients with a WHO stage III/IV (vs. stage I/II) had a 34% decrease in the likelihood of SDI (aRR 0.66; 95% CI: 0.52-0.85) and those in South Africa had a 31% decrease in the likelihood of SDI compared to Zambia (aRR: 0.69; 95% CI: 0.0.60-0.78). We also found that those with TB symptoms had a 16% decrease in the likelihood of SDI (aRR 0.84; 95% CI: 0.73-0.96). Our results also suggest that patients who did have a CD4 count done may have been slightly more likely to initiate on the same day (aRR: 1.09; 95% CI: 0.97-1.22). Sex and age were not predictors of SDI in either model.

## DISCUSSION

In this study, we found that by 2020, uptake of SDI was extensive in Zambia and Malawi, reaching 80-90% of patients initiating or re-initiating ART. It was much less common in South Africa, where just 57% of patients initiated on the same day. Among patients who did not initiate same-day, roughly one third in Malawi, half in South Africa, and nearly all in Zambia went on to initiate within 6 months; the rest had no record of treatment initiation by the end of the 6-month follow-up period.

We found several other publications that reported the rate of same-day initiation in various countries in sub-Saharan Africa, mostly at earlier time periods than our study, which was conducted between 2018 and 2020. A multi-country cohort study using data from the IeDEA Network through 2018, including South Africa but not Malawi or Zambia, reported an overall rate of SDI of 64%[9]. Three studies from South Africa were all completed roughly one year earlier than our study, in 2017-2018. The first, in Johannesburg, reported 20% uptake of same-day initiation[3]. The second, in Johannesburg and Limpopo Province, reported that 40% of patients received SDI, with an increase in uptake from 30.3% at the start of the study period to 54.2% at the end[6]. The third, with data from four provinces, reported 54% SDI uptake in 2018, an increase from 18% in 2016[10]. The difference between these findings and our result of 57% SDI in South Africa in 2018-2020 likely reflects the impact of an additional year of facility experience in implementing the same-day initiation guideline.

In Zambia, uptake of SDI increased from 42% to 75% of newly initiating patients between the beginning of 2016 and the beginning of 2018, a phenomenon attributed largely to the adoption of universal treatment eligibility in 2017[11]. By the time our data were collected in 2018-2020, Zambia’s SDI rate had increased to more than 91% (Table 3). We did not find any published data on SDI uptake in Malawi to compare to our estimate of 88%. Reports from neighboring countries showed SDI rates of 65% in Zimbabwe[7] and 52% in Mozambique in 2017[12]; in Botswana uptake increased from 28% to 59% between 2018 and 2019[13]. SPRINT adds both geographic breadth and, importantly, more recent experience to these prior publications.

Reasons for uptake of SDI or of having delays in ART initiation in our study are not clear. In our data from South Africa and Zambia, only the presence of TB symptoms or a WHO stage of III or IV predicted a delay in initiation. This result likely reflects concerns about starting ART while patients may be experiencing co-infections, such as TB or cryptococcal meningitis. Global guidelines continue to regard these conditions as justification for delaying ART initiation[14], but recent studies suggest that many patients with TB symptoms can and should start ART immediately, as the risk of becoming lost to follow up if ART initiation is delayed may exceed the very low risk of experiencing TB immune reconstitution inflammatory syndrome[15]. Other reasons for delaying initiation may include clinicians’ individual views on SDI, patient preferences (e.g. concerns about stigma, disclosure, or side effects; feeling healthy or not in need of treatment[16]), and/or lack of resolve or resources to adopt new SDI procedures at some facilities.

At the same time, valuable as SDI may be as a strategy for minimizing loss to follow up before starting ART treatment, it is likely that some patients should indeed delay initiation, as they require more urgent medical care before starting ART or have personal concerns about or barriers to treatment that cannot be addressed in a single visit [17,18]. The proportion of patients falling into this latter category is unclear, and it likely varies across different populations. Based on findings of the SLATE II trial in South Africa, a reasonable guess may be that 10-15% of patients are not good candidates for same-day initiation[19]. If this is correct, then both Zambia and Malawi have achieved near-universal uptake of SDI. South Africa still has some distance to go, however, before it has saturated demand for SDI.

The advent of COVID-19 as a major health risk in all three study countries as of the second quarter of 2020 is likely to have changed both patient and provider behavior with regard to ART initiation. On one hand, COVID-19 morbidity, fear of transmission, and societal restrictions kept many potential patients away from the clinic. In South Africa, for example HIV initiations are known to have fallen by 28% in the first year of the pandemic[19]. Conversely, the desire to minimize personal interaction and clinic visits to avoid COVID-19 transmission could have favored same-day initiation for patients who did make it to the clinic to start ART. It is also not clear whether provider and/or patient behavior will revert to earlier norms as pandemic restrictions wane or will sustain any changes that were made. A new round of data collection will be needed to answer those questions.

The SPRINT study had a number of limitations. Our final enrolled sample size was smaller than intended, due to resource limitations that shortened the data collection period. We relied entirely on routinely collected data, which resulted in many missing values for key variables. Despite every effort to enroll a representative sample of patients eligible for ART initiation, we are not confident that all patients who did not initiate ART during the study period were captured in Zambia, potentially leading to overestimates of total initiation in Zambia. Finally, we cannot discern from our data exactly why certain patients faced a delay in starting ART—whether it was a result of patient or provider preference, patient condition, social pressures, or some other barrier. To achieve the full potential of SDI in increasing treatment uptake for those who are eligible and willing—and not creating risks for or coercing those who are not—a better understanding of how decisions are made at healthcare facilities would be of value.

## Data Availability

Data used in the present study are owned by the Ministry of Health or Department of Health in each study country. Requests for data access can be made to the national research authority of each study country.

## Author contributions

AH and SR conceived the study. AH, KH, BP, and TT developed the study protocol and instrument and supervised data collection. AH, KH, and ATB analyzed the data. AH, KH, BP, TT, and SR interpreted the results. AH and SR drafted the manuscript. LM, PH, HS, RN, BWM, and AG provided input on local context, policy, and implementation. All authors approved the final manuscript.

## Funding

This research was supported by the Bill & Melinda Gates Foundation [OPP1192640] to Boston University. The content is solely the responsibility of the authors and does not necessarily represent the official views of the Bill & Melinda Gates Foundation. The funders had no role in study design, data collection and analysis, decision to publish, or preparation of the manuscript.

## Competing interests

LM, RN, and BWM hold positions in government agencies that have supervisory authority over the healthcare facilities involved in this study. No other competing interests were declared.

